# Greater Severity and Functional Impact of Post-Traumatic Headache in Veterans with Comorbid Neck Pain following Traumatic Brain Injury

**DOI:** 10.1101/2020.05.12.20099390

**Authors:** B Shahidi, RW Bursch, JS Carmel, AC Carranza, KM Cooper, JV Lee, CN O’Connor, SF Sorg, KS Maluf, DM Schiehser

## Abstract

**Purpose/Hypothesis:** Traumatic brain injury (TBI) affects 1.7 million people in the U.S. annually, the majority being mild in severity (mTBI). Post-traumatic headache (PTH) is one of the most common symptoms experienced after mTBI caused by head or neck trauma and is often refractory to treatment. Although military Veterans commonly experience mTBI after blast or blunt injuries that likely affect musculoskeletal structures in the neck, the prevalence of cervical symptoms in Veterans with PTH following mTBI have not been well characterized. Similarly, the impact of comorbid neck pain on physical and psychosocial functioning in this population is unknown. This study aims to assess the prevalence of neck pain in Veterans with PTH following mTBI, and to compare the severity and functional impact of PTH between those with and without comorbid neck pain.

**Number of Subjects:** 33 Veterans with PTH after a military-related mTBI were identified from a secondary analysis of data from a prior study.

**Materials and Methods:** Participants were determined to have PTH if they responded to questions on the Patient Headache History Questionnaire (PHHQ), attributed PTH to an accident or injury, and reported PTH in the TBI history. Individuals with both PTH and comorbid neck pain (PTH+NP) were identified based on an affirmative response to one or more questions on the PHHQ indicating that HA episodes were either preceded or accompanied by neck pain. Standardized measures of HA severity and frequency, insomnia, fatigue, mood disorders (Depression, Anxiety, and Posttraumatic Stress Disorder), and physical and emotional role function (SF-36) were compared between groups with and without comorbid neck pain.

**Results:** Of the 33 participants with PTH, 22 (67%) also had neck pain. There were no differences in demographic or TBI-specific characteristics between groups (p>0.069). Sixty-three percent of the PTH+NP group reported severe or incapacitating HA, compared to 27% of those with PTH alone (P=0.049). Insomnia severity and fatigue were significantly greater in the PTH+NP group (P<0.040), and physical function due to bodily pain and physical role function was significantly lower in the PTH+NP group (P<0.036). There were no significant differences between groups for any of the mood or emotional-role functioning scales (P>0.326).

**Conclusions:** The majority of Veterans with mTBI and PTH reported comorbid neck pain. Veterans with PTH and NP reported increased severity of HA, insomnia, fatigue, and a greater physical, but not emotional functional limitations compared to those without NP.

## INTRODUCTION

Approximately 1.7 million people in the United States require medical attention for traumatic brain injury (TBI) every year, with mild TBI (mTBI) being the most prevalent.^1^ Commonly reported symptoms of mTBI include headaches, musculoskeletal pain, fatigue, mood and sleep disturbances, and cognitive impairments. These symptoms are refractory to treatment in nearly half of those who sustain a mTBI, developing into a chronic post-concussive syndrome (PCS) that can persist for years after the traumatic injury^2^. Persistent post-traumatic headache (PTH), defined as a head injury-induced headache lasting greater than 3 months duration,^3^ is the most common symptom of PCS, with prevalence rates averaging 55%-60% across studies.^4,5^

Military service members and veterans are a population with high risk of exposure to TBI, with nearly 40% of those with TBI reporting PTH.^6-8^ In this population, blast-related injuries are the most common cause of TBI due to high exposure to combat and artillery.^9^ One often overlooked feature of blast injury-induced TBI is concurrent damage to surrounding tissues such as cervical musculoskeletal structures that in many ways resembles whiplash injury.^10^ In fact, neck injury is one of the leading causes of medical evacuation of military personnel from US operations.^11^ Although prior literature is sparse on the relationship between neck injury and PTH in the veteran population, the negative impact of concurrent neck injury and headache symptoms has been well established in the civilian population.^12^ For example, the presence of comorbid neck pain with migraine has been shown to be a significant predictor of disability and is associated with increased headache frequency, intensity and duration.^13,14^ In addition, those who experience migraine or PTH report higher rates of concurrent neck pain than those without.^11,13-18^

One clinical barrier to recovery in those with PTH is that high quality evidence to guide specific interventions is lacking. For example, treatments for both PTH and migraine headaches include a wide spectrum of interventions including pharmacological management, nerve blocks, neurostimulation, physical therapy, injections, and even surgery^10,11^. Clinically, PTH often resembles migraine in its presentation, and is therefore managed similarly despite differing mechanisms^19^. As such, it is not surprising that clinical outcomes of PTH treatment are highly variable and suboptimal^11^.

Investigating the influence of comorbid neck pain on physical and emotional functioning among veterans with PTH is necessary to inform clinical management of PTH, and fills an important gap in the literature addressing the military population. In this study, we aim to compare the severity and functional impact of PTH in US Veterans with and without comorbid neck pain following mTBI. We hypothesize that those with PTH and comorbid neck pain will present with greater headache impairments and reduced physical and emotional functioning as compared to those without neck pain.

## METHODS

### Participants

This was a secondary analysis of a database from a prior study^20^. The database included 257 military veterans with and without TBI who were recruited at the VA San Diego Healthcare System (VASDHS) from 2010 to 2014 via outpatient clinics, recruitment flyers, and by word of mouth. The participants included in this database predominantly served in Iraq and Afghanistan conflicts, including Operation Enduring Freedom (OEF), Operation Iraqi Freedom (OIF), and Operation New Dawn (OND). The database contains a compilation of various patient reported questionnaires and neuropsychological outcome measures. Since the current analyses represent a retrospective subsample from a larger study, the sample size was determined based on completion of questionnaires of interest. The VASDHS institutional review board approved the study and all participants provided informed consent.

An eligibility screening was conducted on all participants prior to study enrollment^21^. Participant demographics, as well as relevant military and medical history, were collected in a clinical interview. During this interview, details regarding both military (i.e., active duty) and non-military (i.e., prior to or following military discharge) related head injuries were obtained using a modified version of the VA Semi-Structured Clinical Interview for TBI^22^. For those reporting one or more TBIs, additional information was gathered about the number of TBIs, months since most recent TBI, and mechanism of each TBI injury (i.e., blunt vs. blast). Participants were excluded if they expressed current suicidal and/or homicidal ideation, intent, or plan; had a current or previous diagnosis of a severe mental disorder (e.g. schizophrenia, bipolar disorder); had a history of significant neurological medical conditions such as epilepsy or multiple sclerosis; or reported current substance abuse or dependence as determined by initial diagnostic clinical interview and verified by a Rapid Response 10-drug Test Panel and toxicology screening.

Participants were included in the present study only if they met the following self-report criteria for mild TBI as defined by the Department of Veterans Affairs and Department of Defense^21^: (1) loss of consciousness (LOC) ≤ 30 minutes, (2) alteration of consciousness (AOC) ≤ 24 hours, and/or (3) post-traumatic amnesia (PTA) ≤ 24 hours. Participants were further included if they reported headaches after experiencing a TBI in the clinical interview and completed the Baylor Health Patient Headache History Questionnaire (PHHQ)^23^. This clinical questionnaire collected information on headache severity, frequency, duration, and location, as well as associated neck pain symptoms. Participants in the cohort were split into two groups based on presence (PTH+NP) or absence (PTH) of comorbid neck pain. Individuals with both PTH and comorbid neck pain (PTH+NP) were identified based on an affirmative response to one or more questions on the PHHQ indicating that headache episodes were either preceded or accompanied by neck pain (Figure 1). Patient demographics included age, sex, ethnicity, marital status, years of education, military branch, and employment status.

**Figure 1.**
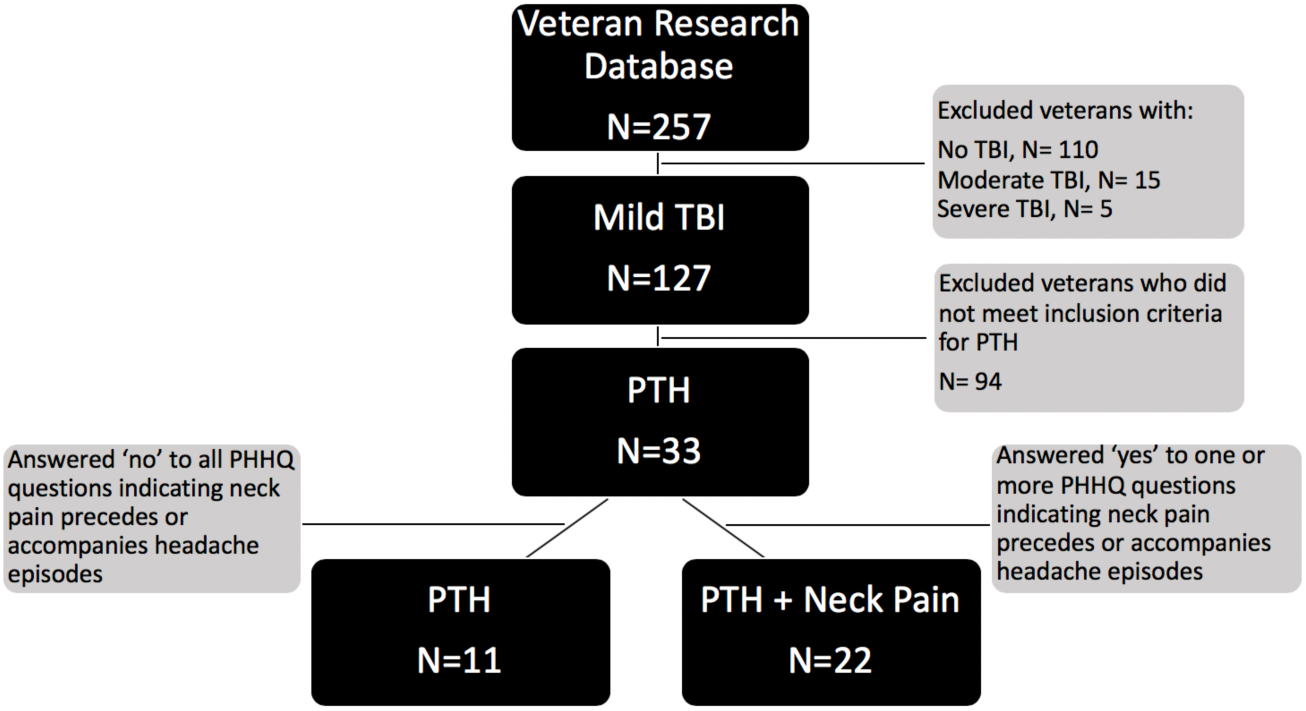
Selection criteria for PTH and PTH+NP study groups sampled from existing Veteran research database. PTH = Post-traumatic headache;’ TBI=Traumatic Brain Injury; PHHQ = Patient Headache History Questionnaire.

### Outcome Measures

#### Headache Impairment Measures

Headache characteristics were assessed using individual survey items from the PHHQ. Due to the small sample size and large number of response options for individual items in this clinical questionnaire, responses were collapsed into dichotomous outcomes for headache severity, frequency, and duration. Three response categories characterized headache location. Severity of headaches was assessed by the PHHQ question “My headaches if I don’t treat them effectively are usually…” with response options of 1) mild or moderate, or 2) severe or incapacitating. Frequency of headaches was assessed by the PHHQ question “My headaches usually occur.” with response options of 1) daily or weekly, or 2) less frequent than weekly. Duration of headaches was assessed by the PHHQ question “My headaches if I don’t take medication will usually last.” with response options of 1) less than 1 day, or 2) more than 1 day. Headache location was assessed by the PHHQ question “My headaches occur.” with response options of 1) only on one side at a time, 2) as a band around my head, or 3) over my entire head.

#### Physical Health Measures

Measures of physical health and function included insomnia, fatigue, general health, bodily pain, physical functioning, and physical role limitations. Sleep was measured using the Insomnia Severity Index (ISI), which is a 7-item questionnaire that quantifies the severity of insomnia with excellent reliability and internal consistency and has been validated in veterans with a history of TBI.^24,25^ ISI scores range from 0-28 pts, with ≥11 points indicating clinically significant insomnia. Fatigue was measured using the Modified Fatigue Impact Scale (MFIS), which is a 29-item questionnaire ranging from 0-84 points that assesses the impact of fatigue on cognitive, physical, and psychosocial functioning and has been validated in veterans with mTBI^26,27^. A threshold of 38 or more points has been suggested to represent clinically significant fatigue^28^. Physical health was measured using physical health subscales of the Short Form Health Survey (SF-36). The SF-36 is a 36-item quality of life survey that determines patient health status with physical health subscales including general health, bodily pain, physical functioning, and physical role limitations, and mental health subscales including mental health, vitality, social functioning, and emotional role limitations. Each of the 8 subscales are scored separately and range from 0-100, with higher scores indicating better health and lower scores indicating more limitation.^29^

#### Emotional Health Measures

Emotional health metrics assessed depression, anxiety, post-traumatic stress, mental health, vitality, social functioning, and emotional role limitations. Depression and anxiety were assessed using the Beck Depression Inventory (BDI) and Beck Anxiety Inventory (BAI), respectively. The BDI is a 21-item self-report questionnaire with good reliability and validity that is scored on a scale from 0-63 points. The BDI has been recommended for evaluation of depression in the TBI population.^30,31^ Established score ranges for interpreting the severity of depression are as follows; 17-20 points indicates borderline clinical depression, 21-30 points indicates moderate depression, and over 40 points indicates severe depression.^32^ The BAI is a 21-item self-report questionnaire with excellent reliability and validity.^33^ It is scored on a scale of 0-63 points, with scores over 36 points indicating high levels of anxiety, and scores below 21 points indicating low anxiety.^33^ Presence and severity of Post-traumatic Stress Disorder (PTSD) were measured using the Post-traumatic Stress Disorder Checklist (PCL) Military Version. The PCL is a 17-item self-report measure that is scored on a scale from 0-85 points, with scores above 50 points indicating clinically significant PTSD^34^. Mental health, vitality, social functioning, and emotional role limitations were calculated from the mental health subscales of the SF-36 as described above.

### Statistical Analysis

Independent t-tests were used to compare means between PTH and PTH+NP groups for continuous variables. Chi square or fisher exact tests were used to compare proportions between groups for categorical variables. P values of 0.05 were considered significant, and P values of less than 0.08 were considered a trend. Data were examined for outliers and checked for normality. All statistical analyses were performed on SPSS software version 25 (IBM Corp., 2019).

## RESULTS

### Participant Characteristics

A total of 33 participants met the inclusion criteria for this study, with the majority (N=22, 66.7%) reporting concurrent neck pain. Participant characteristics are detailed in Table 1. The majority of participants were Caucasian (57.6%), with no significant difference in the ethnic distribution between groups (P=0.196).

**Table 1.**
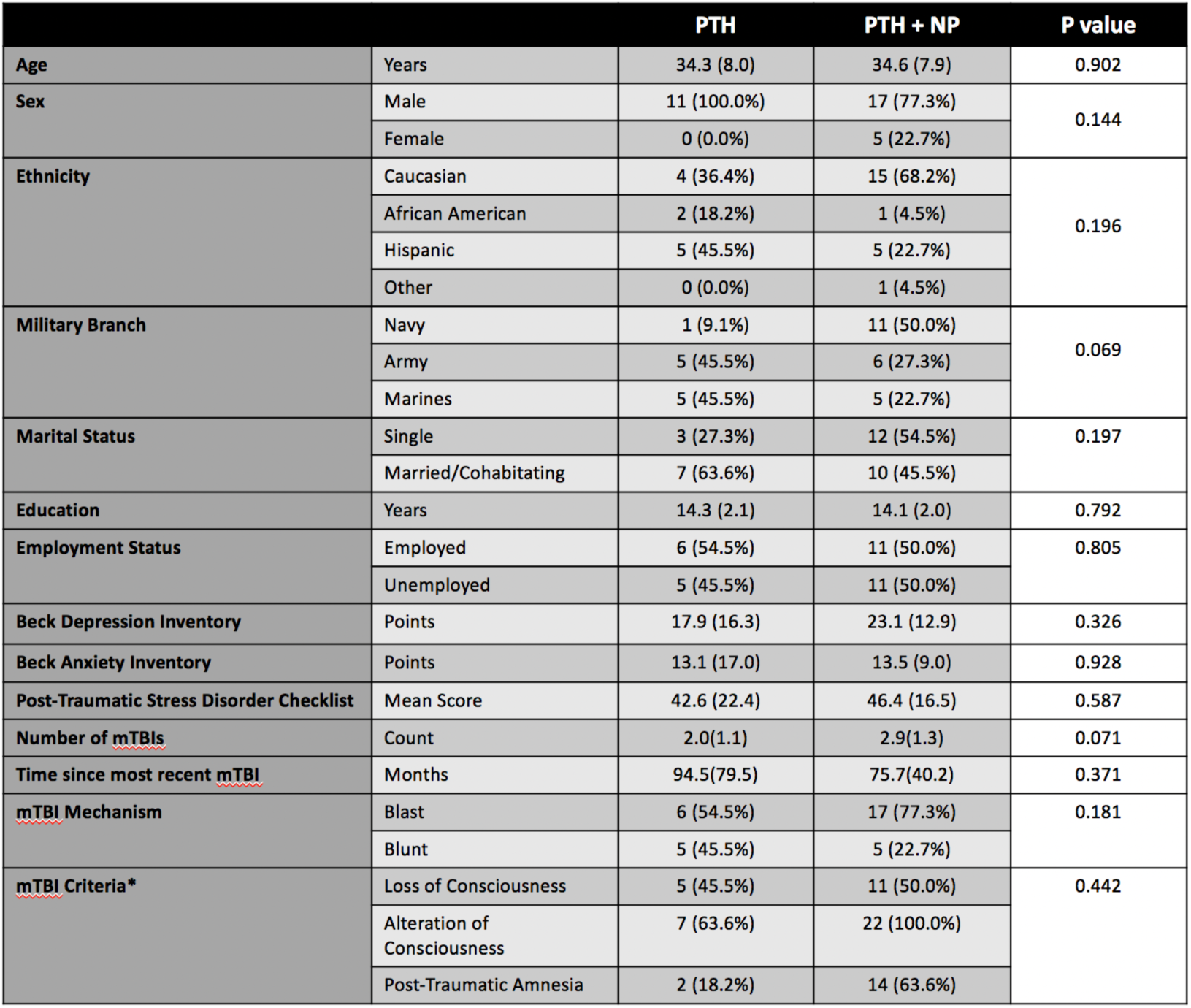
Participant demographic and clinical characteristics. Values reported as mean (SD) or N (% group total). PTH= Post-traumatic headache, NP = Neck Pain. individuals may have experienced multiple mTBI criteria simultaneously.

|  |  | PTH | PTH + NP | P value |
| --- | --- | --- | --- | --- |
| Age | Years | 34.3 (8.0) | 34.6 (7.9) | 0.902 |
| Sex | Male | 11 (100.0%) | 17 (77.3%) | 0.144 |
|  | Female | 0 (0.0%) | 5 (22.7%) |  |
| Ethnicity | Caucasian | 4 (36.4%) | 15 (68.2%) | 0.196 |
|  | African American | 2 (18.2%) | 1 (4.5%) |  |
|  | Hispanic | 5 (45.5%) | 5 (22.7%) |  |
|  | Other | 0 (0.0%) | 1 (4.5%) |  |
| Military Branch | Navy | 1 (9.1%) | 11 (50.0%) | 0.069 |
|  | Army | 5 (45.5%) | 6 (27.3%) |  |
|  | Marines | 5 (45.5%) | 5 (22.7%) |  |
| Marital Status | Single | 3 (27.3%) | 12 (54.5%) | 0.197 |
|  | Married/Cohabiting | 7 (63.6%) | 10 (45.5%) |  |
| Education | Years | 14.3 (2.1) | 14.1 (2.0) | 0.792 |
| Employment Status | Employed | 6 (54.5%) | 11 (50.0%) | 0.805 |
|  | Unemployed | 5 (45.5%) | 11 (50.0%) |  |
| Beck Depression Inventory | Points | 17.9 (16.3) | 23.1 (12.9) | 0.326 |
| Beck Anxiety Inventory | Points | 13.1 (17.0) | 13.5 (9.0) | 0.928 |
| Post-Traumatic Stress Disorder Checklist | Mean Score | 42.6 (22.4) | 46.4 (16.5) | 0.587 |
| Number of mTBIs | Count | 2.0(1.1) | 2.9(1.3) | 0.071 |
| Time since most recent mTBI | Months | 94.5(79.5) | 75.7(40.2) | 0.371 |
| mTBI Mechanism | Blast | 6 (54.5%) | 17 (77.3%) | 0.181 |
|  | Blunt | 5 (45.5%) | 5 (22.7%) |  |
| mTBI Criteria* | Loss of Consciousness | 5 (45.5%) | 11 (50.0%) | 0.442 |
|  | Alteration of Consciousness | 7 (63.6%) | 22 (100.0%) |  |
|  | Post-Traumatic Amnesia | 2 (18.2%) | 14 (63.6%) |  |

The veteran cohort was predominantly male (84.8%), with no significant difference in sex between groups (P = 0.144). The PTH+NP group tended to have more Navy veterans as compared to the PTH group (P=0.069). Just over half of the cohort was employed (51.5%), with no differences in marital status (P=0.197), education (P=0.792), or employment status between groups (P = 0.805). The PTH+NP group tended to report a greater number of mTBIs compared to the PTN group (P = 0.071), with no differences in time since injury, injury mechanism, or mTBI diagnostic criteria characteristics (P ≥ 0.181).

### Headache Impairments

Headache severity was greater in the PTH+NP group than the PTH group (P = 0.049), with the majority of participants with concurrent neck pain reporting severe or incapacitating headaches and those without headaches reporting only mild or moderate headaches. There were no differences in headache duration or frequency between groups (P ≥ 0.231), and no clear pattern emerged for differences in headache location between those with and without concurrent neck pain (Table 2).

**Table 2.**
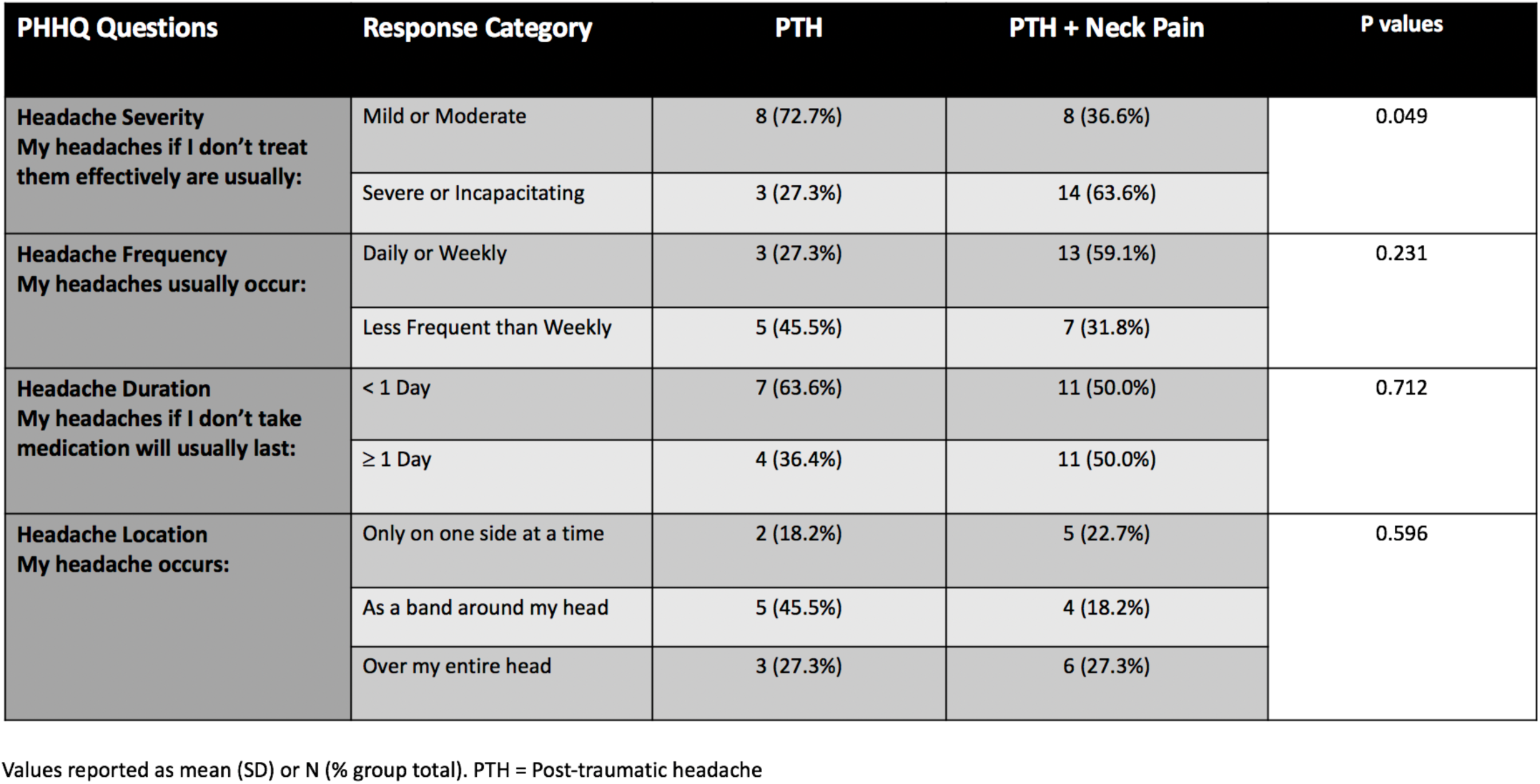
Headache characteristics in participants with and without concurrent neck pain

| PHHQ Questions | Response Category | PTH | PTH + Neck Pain | P values |
| --- | --- | --- | --- | --- |
| Headache Severity<br>My headaches if I don't treat them effectively are usually: | Mild or Moderate | 8 (72.7%) | 8 (36.6%) | 0.049 |
|  | Severe or Incapacitating | 3 (27.3%) | 14 (63.6%) |  |
| Headache Frequency<br>My headaches usually occur: | Daily or Weekly | 3 (27.3%) | 13 (59.1%) | 0.231 |
|  | Less Frequent than Weekly | 5 (45.5%) | 7 (31.8%) |  |
| Headache Duration<br>My headaches if I don't take medication will usually last: | < 1 Day | 7 (63.6%) | 11 (50.0%) | 0.712 |
| | $\geq 1$ Day | 4 (36.4%) | 11 (50.0%) | |
| Headache Location<br>My headache occurs: | Only on one side at a time | 2 (18.2%) | 5 (22.7%) | 0.596 |
|  | As a band around my head | 5 (45.5%) | 4 (18.2%) |  |
|  | Over my entire head | 3 (27.3%) | 6 (27.3%) |  |

Values reported as mean (SD) or N (% group total). PTH = Post-traumatic headache

### Physical Health Measures

Severity of insomnia was higher in the PTH+NP group with those reporting comorbid neck pain indicating clinically relevant insomnia of (17.5 ± 6.3), as compared to the PTH+NP group (9.4 ± 7.6; P = 0.003, Figure 2A). On average, fatigue levels in the PTH+NP group exceeded the threshold for clinically relevant fatigue (44.8 ± 22.3) and were significantly higher than the PTH group (21.2 ± 35.7; P = 0.040, Figure 2B). The PTH+NP group reported lower physical function due to bodily pain (42.2 ± 24.2) than the PTH group (67.7 ± 28.8; P = 0.012, Figure 2C), and lower physical-health related role function (27.3 ± 38.5) than the PTH group (59.1 ± 40.7; P = 0.036, Figure 2D). SF-36 general health (P = 0.389) did not differ between groups, and there was a trend for reduced physical functioning (P = 0.084) in the PTH+NP group.

**Figure 2:**
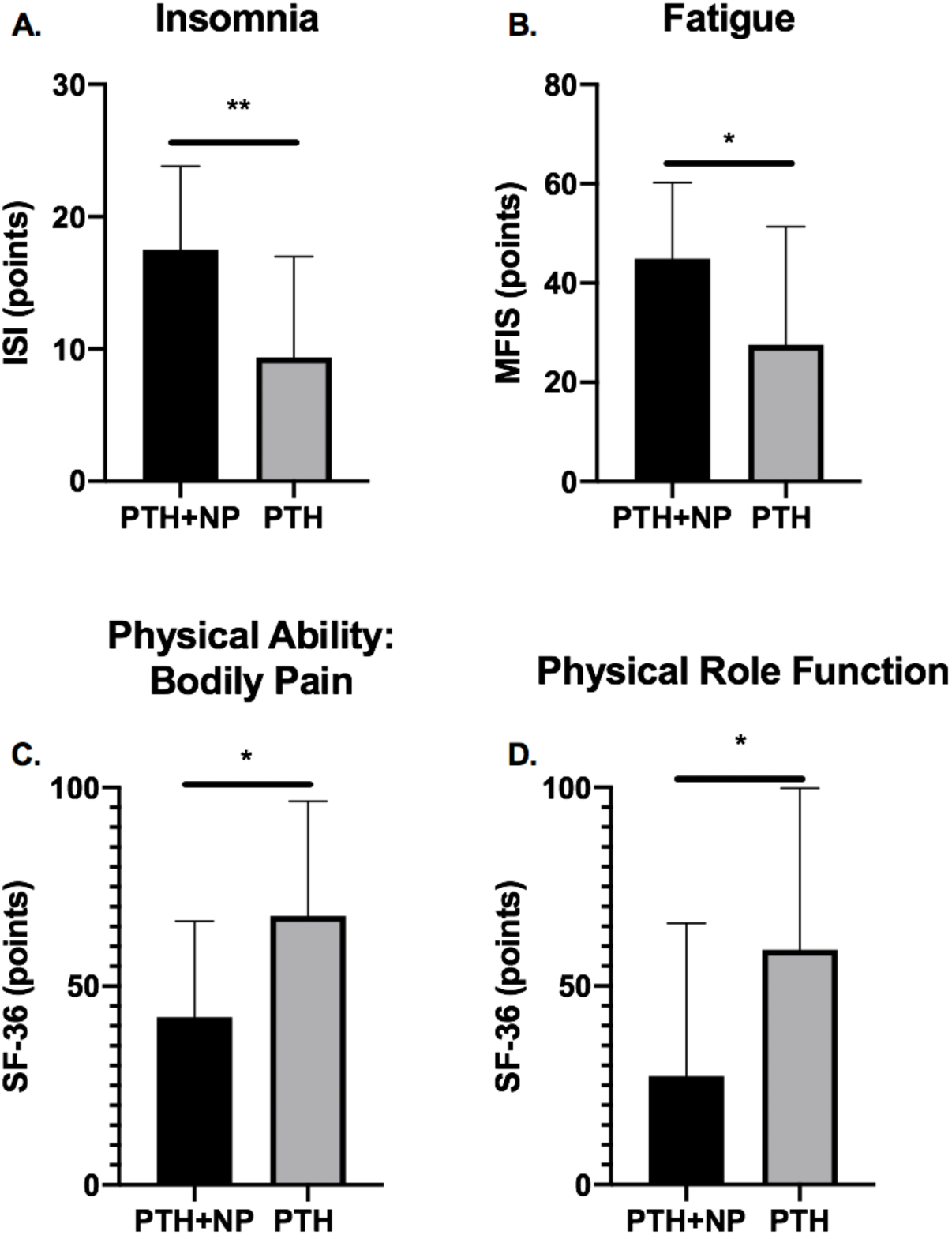
Bars indicate mean (SD) scores for the post-traumatic headache with neck pain (PTH+NP, N=22) and PTH with no neck pain (PTH, N=11) groups. A. Insomnia Severity Index (ISI) scores range from 0-28, with higher scores indicating increased insomnia severity. B. Modified Fatigue Impact Scale Scores range from 0-84, with higher scores indicating increased fatigue. C. SF-36 Bodily Pain domain scores, with lower scores indicating lower physical ability due to bodily pain. D. SF-36 Physical Role Function domain scores, with lower scores indicating lower physical role functioning. (**P<0.01, *P<0.05)

### Emotional Health Measures

There were no significant differences in mood-related measures of depression (BDI, P = 0.326), anxiety (BAI, P = 0.928), or PTSD (PCL, P = 0.587) between groups. Similarly, there were no group differences in the SF-36 Mental Health subscale (PTH+NP = 50.2 ± 19.4; PTH = 56.7 ± 26.9; P = 0.429; Figure 3A), or role limitations related to emotional health as measured by the SF-36 Role Emotional subscale (PTH+NP = 36.4 ± 41.0; PTH = 51.5 ± 45.6; P = 0.343, Figure 3B). SF-36 Vitality (P = 0.389) and Social Functioning (P = 0.985) subscale scores were also similar between groups.

**Figure 3.**
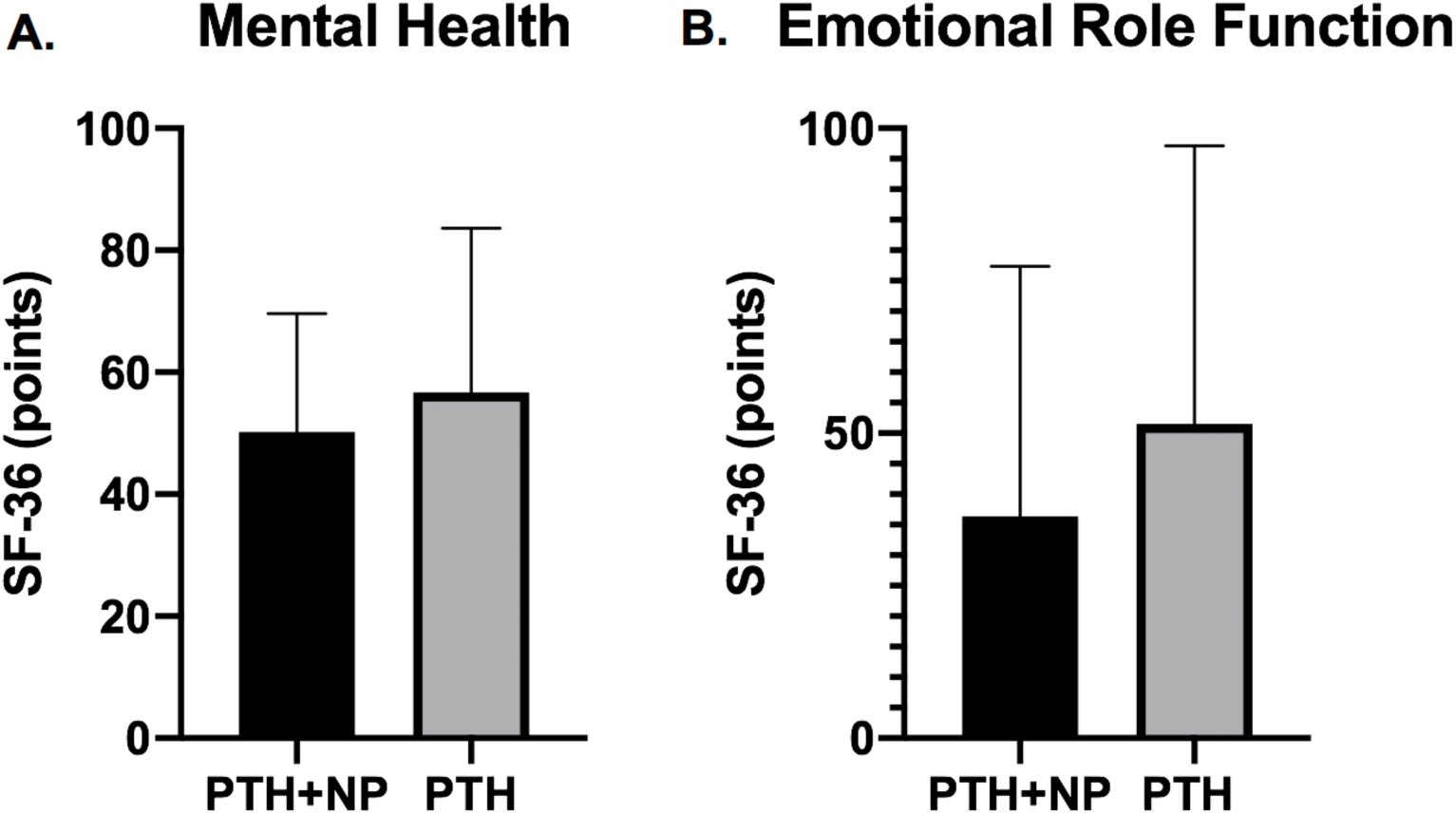
Bars indicate mean (SD) scores for the post-traumatic headache with neck pain (PTH+NP, N=22) and PTH with no neck pain (PTH, N=11) groups. A. SF-36 Mental Health domain scores, with lower scores indicating lower mental health. B. SF-36 Emotional Role domain scores, with lower scores indicating lower emotional role functioning. All scores range from 0-100 points, with no significant differences between groups (P>0.05).

## DISCUSSION

The majority of veterans with mTBI and PTH in our sample experienced concurrent neck pain that was associated with more severe headaches, insomnia, and fatigue, as well as greater physical limitations due to bodily pain and reduced physical role functioning compared to those without neck pain. Importantly, these physical limitations occurred despite similar mood profiles and emotional functioning between groups. Overall, these findings support our hypothesis that veterans with PTH and comorbid neck pain experience more severe headaches and subsequent physical limitations, but are contrary to our hypothesis regarding the relationship between comorbid neck pain and emotional functioning. This is the first study to show that comorbid neck pain negatively impacts headache symptoms and physical role functioning in veterans with PTH following mTBI.

Despite the limited size of our cohort, the prevalence rate of 66.7% for comorbid neck pain is consistent with prevalence rates for neck pain reported in larger samples. One paper investigating the prevalence of chronic musculoskeletal pain in civilians after mTBI reported that 59.6% of individuals reported neck or shoulder pain of greater than 6 month duration^35^, and another study reported a neck pain prevalence rate of 41.7% in military veterans^4^. These rates mirror the prevalence of comorbid neck in primary headache disorders such as migraine, where prevalence rates are reported to range between 40%-76%.^13,14,16^ To our knowledge, no previous study reporting neck pain prevalence rates in individuals with mTBI has compared health or function between those with and without this common comorbidity.

The relationship between neck pain and headache impairments has not been well elucidated in the TBI population. Existing studies suggest that the majority of individuals with mTBI experience headaches on a daily basis, and headache severities are moderate, with average numerical pain rating scores ranging from 4 to 6 on a 10-point scale.^4,35^ Our data indicate that the majority of individuals with comorbid neck pain reported severe or incapacitating headaches compared to only mild or moderate severity in those without, suggesting that neck pain significantly compounds headache symptom intensity in this population. Despite this difference in symptom intensity, we found no influence of neck pain on headache frequency or duration, which is in contrast to studies of migraine reporting positive correlations between neck pain or neck pain related disability and headache frequency^13,14,36^

One interesting finding of our study was that the location of headache was non-specific in our PTH population and did not differ between groups. Primary and cervicogenic headaches are often closely tied to neck pain. Cervicogenic headache typically presents with unilateral symptoms and is by definition caused by a disorder of the cervical spine and its component bony, disc and/or soft tissue elements.^3,37^ A similar presentation in the PTH+NP group would suggest that similar treatment approaches targeting cervical pain generators may be indicated for both PTH and cervicogenic headaches. However, the variation in phenotype observed here suggests the need for a different clinical approach, and perhaps supports the current clinical practice to treat PTH similarly to migraine headaches which are thought to be centrally mediated. Elevated fatigue and insomnia in the PTH+NP group support the hypothesis that comorbid neck pain may result from heightened central sensitivity in some individuals with mTBI and PTH^38^. Although there is some evidence that impaired descending pain modulation may contribute to PTH^39^, future studies are needed to identify the relative contribution of peripheral and central mechanisms to greater symptom severity among those with PTH and comorbid neck.

Where the influence of comorbid neck pain on headache impairments seems to be variable-dependent, its influence on overall physical health and ability is clear, and mirrors findings from studies in other headache populations. In the TBI population, sleep disturbances are common; occurring in nearly half of individuals, with effects lasting from months to years after the original injury and increasing with injury severity.^40-46^ Similar relationships exist in individuals with persistent headaches^47,48^, and even chronic neck pain.^49^ As such, it is not surprising that our results reflect that a combination of mTBI, headache, and neck pain produces a more severe phenotype than any given symptom in isolation. Closely related to insomnia, fatigue is another symptom strongly established among those with mTBI that can persist for months or years following the injury.^45,50,51^ Fatigue has been shown to be a strong predictor for disability in the TBI population.^52^ Moreover, those with neck pain of any source tend to report higher levels of fatigue when compared to their asymptomatic counterparts.^53^ These relationships also extend to overall disability, although the literature is mixed in this regard. The presence of neck pain has been shown to be significantly associated with physical impairments (mobility and strength)^54^ as well as disability in individuals with migraine headaches,^14,36^ while other research has failed to demonstrate a significant influence on participation restrictions such as missing days of work or social events.^54^ Although these data are largely limited to populations that are unrelated to mechanisms including trauma, the trend in the literature towards increased physical disability among those with neck pain coincides with our findings of overall reduced ability on the SF-36 physical health subscales.

Interestingly, our findings related to mood and emotional health did not distinguish between groups. This was surprising given the tendency for a greater number of head injuries in the PTH+NP group, which has previously been shown to correlate with the severity of mood disturbances^55^. This finding was also contrary to our initial hypothesis given that psychological and emotional health impairments are widely observed in the mTBI population^20^, as well as in those with headaches and neck pain^56-59^. Despite the lack of group differences, our data show that both groups demonstrated clinically relevant levels of anxiety and depression, as well as reduced mental health and emotional role functioning. The similarity of mood and emotional health ratings between groups suggest that differences observed in headache severity and physical health domains were not confounded by differences in emotional health or a tendency of the PTH+NP group to endorse greater distress generally. The lack of a direct relationship between psychological and physical symptomology has been recognized in recent literature in the mTBI population, suggesting that these factors may be independently modulated^5,60^.

### Study Limitations

This study had a number of limitations, primarily that the sample size was small due to the secondary nature of our analysis. Despite the small size of this sample, it is comparable to other studies of PTH in mTBI populations evaluating impairment-based measures and physical health, and the ability to distinguish statistical differences between groups for a number of our outcomes suggests sufficient power for our comparisons of interest. Additionally, our retrospective analysis of an existing database resulted in a lack of prospectively defined variables of interest. As such, our operational definition of neck pain was based on a small set of questions from the PHHQ, which limited our ability to characterize the nature and severity of neck pain symptoms. Additionally, causal inferences cannot be determined from a cross-sectional study design. Future research is needed to prospectively evaluate the mechanisms and impact of comorbid neck pain on headache impairments, physical, and emotional health in this population.

## CONCLUSION

Neck pain is a common comorbidity of PTH and one that has several functional implications. In this sample of veterans with mTBI and PTH, those with comorbid neck pain experienced greater headache severity and poorer physical health and role functioning, despite no differences in emotional health compared to those without comorbid neck pain. Awareness of these associated impairments may be a valuable contribution to direct future research and clinical practices. Further research that prospectively investigates the mechanisms and impact of comorbid neck pain on headache impairments, physical and psychosocial functioning is needed in this population, specifically with regard to directing targeted treatment approaches.

## Data Availability

N/A

## Notes

### Competing Interest Statement

The authors have declared no competing interest.

